# Exploring the causal role of the human gut microbiome in endometrial cancer: a Mendelian randomization approach

**DOI:** 10.1101/2024.03.06.24303765

**Authors:** Ella Fryer, Charlie Hatcher, Rochelle Knight, Kaitlin Wade

**Affiliations:** MRC Integrative Epidemiology Unit, University of Bristol, Oakfield House, Oakfield Grove, Bristol, BS8 2BN, UK; Population Health Sciences, Bristol Medical School, University of Bristol, Bristol, BS8 2BN, UK

**Keywords:** Gut microbiome, endometrial cancer, Mendelian randomization

## Abstract

Endometrial cancer presents a major public health issue, particularly in post-menopausal women. Whilst there are known risk factors for the disease, including oestrogen and obesity, these factors do not fully explain risk variability in cancer outcomes. The identification of novel risk factors may aid in better understanding of endometrial cancer development and, given the link with oestrogen metabolism, obesity and the risk of various cancers, the gut microbiome could be one such risk factor. Mendelian randomization (MR), a method that reduces biases of conventional epidemiological studies (namely, confounding and reverse causation) by using genetic variants to proxy exposures, was used to investigate the effect of gut microbial traits on endometrial cancer risk. Whilst our initial analyses showed that the presence of an unclassified group of bacteria in the *Erysipelotrichaceae* family increased the risk of oestrogen-dependent endometrial cancer (odds ratio (OR) per approximate doubling of the genetic liability to presence vs absence: 1.13; 95% CI: 1.01, 1.26; P=0.03), subsequent sensitivity analyses, including colocalisation, suggested these findings were unlikely reflective of causality. This work highlights the importance of using a robust MR analysis pipeline, including sensitivity analyses to assess the validity of causal effect estimates obtained using MR.

## INTRODUCTION

Endometrial cancer is a major public health issue, being the fourth most common cancer in women in the UK and US and the sixth most common cancer worldwide (1, 2). In 2023, there were an estimated 66,200 new cases in the US, which was 3.4% of all new cancer cases, and an estimated 13,030 deaths (3). There are two main subtypes of endometrial cancer, endometrioid (or oestrogen-dependent endometrial cancer) and non-endometrioid. More than 70% of endometrial cancer cases are endometrioid and occur in women over 50 (1, 4). Targeting this risk group with novel interventions could be a key strategy to preventing disease occurence and reducing overall disease burden.

There are a number of known risk factors for endometrioid cancer, including obesity, insulin resistance and excess exposure to oestrogen (1). However, the mechanisms by which these risk factors interact and affect cancer development is not fully understood and current research is unable to explain the varying cancer outcomes observed in high-risk groups (5). One explanation may be that there are novel risk factors that have not been discovered and, therefore, the relationship between these risk factors and endometrial cancer is yet to be quantified. The gut microbiome may be one such risk factor, as previous observational studies have linked the gut microbiome with many other cancers, including colorectal and breast cancers (6). The gut microbiome is also thought to modulate obesity and oestrogen metabolism, which could explain how it may play a causal role in the development of endometrial cancer (5, 7).

Previous studies have investigated the role of the gut microbiome in endometrial cancer development using in vivo and in vitro experiments as well as observational studies in humans using small numbers of cancer cases and controls. Results from these studies have found altered gut microbial profiles in cancer cases compared to controls. However, many of these studies are likely to suffer limitations such as reverse causation (i.e., where the gut microbiome may be a consequence of endometrial cancer diagnosis rather than a cause) or confounding (i.e., whereby variation in the gut microbiome is altered independently by the causal risk factor for endometrial cancer, but no true relationship exists between the gut microbiome and cancer outcomes).

Mendelian randomization (MR) is a method that uses germline genetic variants that are robustly associated with a trait of interest (e.g., the gut microbiome) as a proxy for that trait to improve causal inference in the relationship between that trait and an outcome (e.g., endometrial cancer) (8, 9). As germline variants are fixed and randomly allocated at conception, using these variants as proxies for exposures has a number of advantages, including removing the possibility of reverse causation and reducing phenotypic confounding that may distort observed relationships. MR is therefore a useful alternative to randomised control trials (RCTs), which are often not feasible, due to financial, practical and ethical considerations.

Therefore, we aimed to use two-sample MR, which utilises summary-level data, to estimate the causal relationship between the gut microbiome and endometrial cancer using some of the largest genome-wide association studies (GWASs) to date and, importantly, a series of sensitivity analyses that tested the robustness of findings to violations of MR assumptions.

## RESULTS

### Two-sample MR analyses

To determine whether the gut microbiome plays a causal role in endometrial cancer, we performed two-sample MR. Summary-level data was obtained from a recent microbiome GWAS (mGWAS) meta-analysis (n=3,890) (10) and a GWAS meta-analysis of endometrial cancer (cases=12,906, controls=108,979), including 8,758 endometrioid cancer cases and 1,230 non-endometrioid cancer cases (11), both conducted in non-overlapping populations of European ancestry. There were 13 single nucleotide polymorphisms (SNPs) in the mGWAS that exceeded a genome-wide p-value threshold (p<5×10^-08^), each associated with an independent microbial trait, as well as an additional SNP that has been reliably associated with bacteria in the *Bifidobacterium* genus across multiple mGWASs (Supplementary Table 1) (12–14). These 14 SNPs were extracted from the endometrial cancer GWAS summary statistics from the Epidemiology of Endometrial Cancer Consortium (E2C2) consortium, which were available for different histological subtypes in the IEU OpenGWAS (GWAS IDs: ebi-a-GCST006464-6) (15). Several of the SNPs were not present in the endometrial cancer data, or only present for some subtypes, and it was not possible to identify a proxy SNP, the corresponding exposures were therefore excluded from subsequent analyses.

Two-sample MR analyses found evidence that the presence (vs absence) of an unclassified group of bacteria in the *Erysipelotrichaceae* family (*G. unclassified, F. Erysipelotrichaceae*) increased the risk of endometrioid cancer by 13% (odds ratio (OR) per approximate doubling of the genetic liability to presence vs absence: 1.13; 95% CI: 1.01, 1.26; P=0.03). When including all histologies in the analysis, the effect estimate for this microbial trait was in the same direction and of a similar magnitude, however the confidence intervals (CIs) crossed the null (OR: 1.08; 95% CI: 0.99, 1.19; P=0.09). In non-endometrioid cancer, the effect estimate for this microbial trait was in the opposite direction; however, CIs crossed the null (OR: 0.94; 95% CI: 0.72, 1.22; P=0.64). MR effect estimates for the remaining microbial traits on all histologies and subtypes of endometrial cancer were smaller and CIs crossed the null (Table 1, Figure 1). Thus, all sensitivity analyses were based on the effects observed between *G. unclassified, F. Erysipelotrichaceae* and endometrioid cancer only.

**Figure 1.**
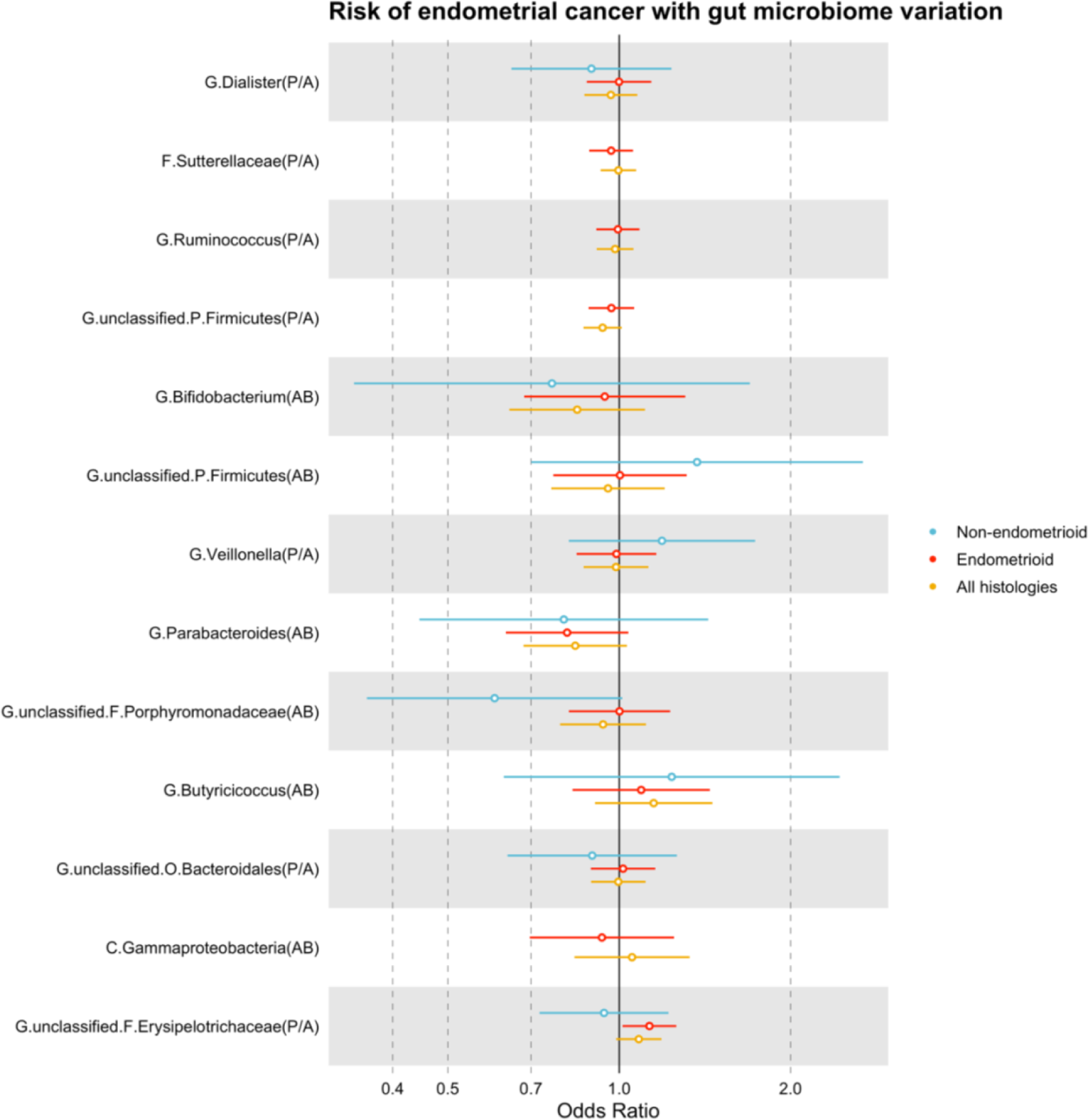
MR estimates of the effect of each microbial trait on endometrial cancer and subtypes. Abbreviations: AB = abundance; CI = confidence interval; MR = Mendelian randomization; OR = odds ratio; P/A = presence vs absence; SD = standard deviation. MR estimates represent the OR for endometrial risk and 95% CI per SD unit change for continuous microbial traits (AB) or per approximate doubling of the genetic liability to presence (vs absence) of each binary microbial trait (P/A).

**Table 1.**
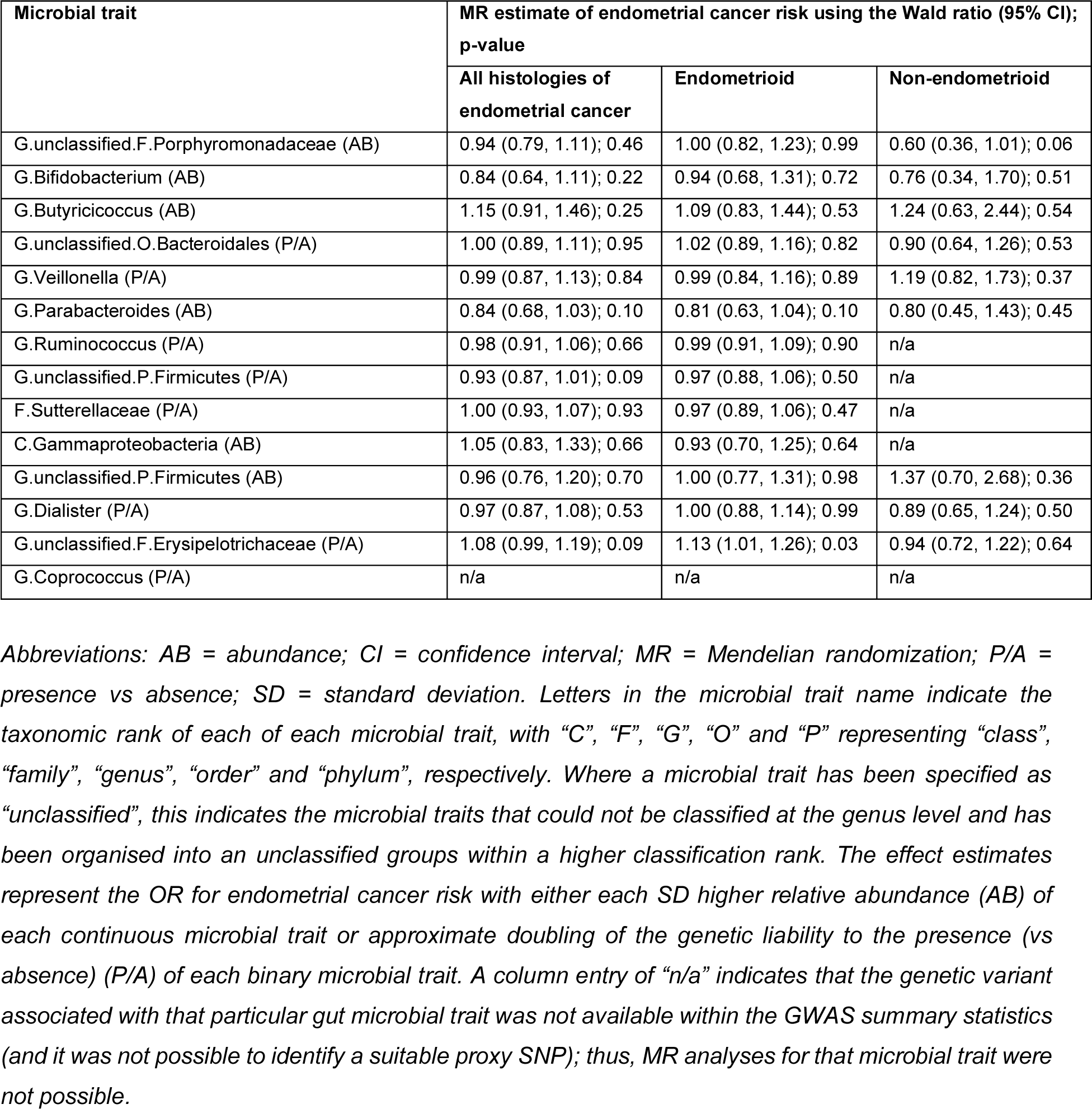
Two-sample MR estimates of the effect of each microbial trait on endometrial cancer and both endometrioid and non-endometrioid subtypes.

| Microbial trait | MR estimate of endometrial cancer risk using the Wald ratio (95% CI); p-value |  |  |
| --- | --- | --- | --- |
|  | All histologies of endometrial cancer | Endometrioid | Non-endometrioid |
| G.unclassified.F.Porphyromonadaceae (AB) | 0.94 (0.79, 1.11); 0.46 | 1.00 (0.82, 1.23); 0.99 | 0.60 (0.36, 1.01); 0.06 |
| G.Bifidobacterium (AB) | 0.84 (0.64, 1.11); 0.22 | 0.94 (0.68, 1.31); 0.72 | 0.76 (0.34, 1.70); 0.51 |
| G.Butyricicoccus (AB) | 1.15 (0.91, 1.46); 0.25 | 1.09 (0.83, 1.44); 0.53 | 1.24 (0.63, 2.44); 0.54 |
| G.unclassified.O.Bacteroidales (P/A) | 1.00 (0.89, 1.11); 0.95 | 1.02 (0.89, 1.16); 0.82 | 0.90 (0.64, 1.26); 0.53 |
| G.Veillonella (P/A) | 0.99 (0.87, 1.13); 0.84 | 0.99 (0.84, 1.16); 0.89 | 1.19 (0.82, 1.73); 0.37 |
| G.Parabacteroides (AB) | 0.84 (0.68, 1.03); 0.10 | 0.81 (0.63, 1.04); 0.10 | 0.80 (0.45, 1.43); 0.45 |
| G.Ruminococcus (P/A) | 0.98 (0.91, 1.06); 0.66 | 0.99 (0.91, 1.09); 0.90 | n/a |
| G.unclassified.P.Firmicutes (P/A) | 0.93 (0.87, 1.01); 0.09 | 0.97 (0.88, 1.06); 0.50 | n/a |
| F.Sutterellaceae (P/A) | 1.00 (0.93, 1.07); 0.93 | 0.97 (0.89, 1.06); 0.47 | n/a |
| C.Gammaproteobacteria (AB) | 1.05 (0.83, 1.33); 0.66 | 0.93 (0.70, 1.25); 0.64 | n/a |
| G.unclassified.P.Firmicutes (AB) | 0.96 (0.76, 1.20); 0.70 | 1.00 (0.77, 1.31); 0.98 | 1.37 (0.70, 2.68); 0.36 |
| G.Dialister (P/A) | 0.97 (0.87, 1.08); 0.53 | 1.00 (0.88, 1.14); 0.99 | 0.89 (0.65, 1.24); 0.50 |
| G.unclassified.F.Erysipelotrichaceae (P/A) | 1.08 (0.99, 1.19); 0.09 | 1.13 (1.01, 1.26); 0.03 | 0.94 (0.72, 1.22); 0.64 |
| G.Coprococcus (P/A) | n/a | n/a | n/a |

*Abbreviations: AB = abundance; CI = confidence interval; MR = Mendelian randomization; P/A = presence vs absence; SD = standard deviation. Letters in the microbial trait name indicate the taxonomic rank of each of each microbial trait, with “C”, “F”, “G”, “O” and “P” representing “class”, “family”, “genus”, “order” and “phylum”, respectively. Where a microbial trait has been specified as “unclassified”, this indicates the microbial traits that could not be classified at the genus level and has been organised into an unclassified groups within a higher classification rank. The effect estimates represent the OR for endometrial cancer risk with either each SD higher relative abundance (AB) of each continuous microbial trait or approximate doubling of the genetic liability to the presence (vs absence) (P/A) of each binary microbial trait. A column entry of “n/a” indicates that the genetic variant associated with that particular gut microbial trait was not available within the GWAS summary statistics (and it was not possible to identify a suitable proxy SNP); thus, MR analyses for that microbial trait were not possible.*

### Sensitivity analyses

#### Manual exploration of pleiotropy

We performed several sensitivity analyses to test for violations of MR assumptions and explore other explanations for the observed causal effect of *G. unclassified, F. Erysipelotrichaceae* on endometrioid cancer. We were unable to perform formal methods to test for horizontal pleiotropy, owing to the fact that only one SNP was available to proxy for each microbial trait. We therefore searched the IEU OpenGWAS (15) and PhenoScanner (16) platforms for the SNP associated with *G. unclassified, F. Erysipelotrichaceae* (rs6733298) to test whether it had previously been associated with endometrial cancer or any other traits that may influence endometrial cancer independently from this gut microbial trait. A lenient multiple testing p-value threshold (P<1×10^−04^) was used to filter associations. In the IEU OpenGWAS, there were a total of seven SNP-trait associations that met this threshold; these traits were leg fat percentage (left and right), leg fat mass (left and right), body fat percentage, educational qualifications and years of schooling (Supplementary Table 2). In PhenoScanner, two SNP-trait associations met this threshold, including long-standing illness or disability and coagulation defects (Supplementary Table 3). Some of the associations just below the multiple testing threshold were associated with increased leg fat percentage. This SNP had also previously been associated with 48 expression quantitative traits (eQTs) and 12 methylation quantitative traits (mQTs) (Supplementary Table 4 and 5), but we found no evidence for associations with proteins or metabolites (Supplementary Table 6 and 7). Given the association of this SNP with complex traits with biological relevance to endometrioid cancer (17), there may be a likely alternative causal pathway between this SNP and endometrioid cancer that is independent of *G. unclassified, F. Erysipelotrichaceae,* providing some evidence for horizontal pleiotropy. However, given the relationship between the gut microbiome and adiposity-related traits and the uncertainty of the direction of this association, it is difficult to unpick whether this reflects horizontal pleiotropy (i.e., bias inducing) or vertical pleiotropy (i.e., non-bias inducing and that which is part of the causal pathway between the gut microbiome and endometrial cancer).

#### Two-sample MR using a lenient p-value threshold for selection of genetic instruments

In the original MR analyses, only one SNP was associated with *G. unclassified, F. Erysipelotrichaceae*. In order to increase power and test for the possibility that this effect was driven by horizontal pleiotropy, a more lenient p-value threshold (p<1×10^-05^) was used to select instruments for this microbial trait and estimate its causal effecton endometrioid cancer. After restricting this greater set of SNPs to those that had a consistent effect estimate across the cohorts included in the original mGWAS, four SNPs were available to use as instruments to proxy *G. unclassified, F. Erysipelotrichaceae* (Supplementary Table 8). In the subsequent MR analyses, the inverse-variance weighted(IVW)-derived effect estimate of this microbial trait on endometrioid cancer was found to be consistent in direction to the estimate obtained using the Wald ratio in the main analyses; however, the magnitude was attenuated and the CIs spanned the null (OR: 1.02; 95% CI: 0.93, 1.11; P=0.67) (Supplementary Figure 1).

Estimates of the causal effect of the presence of *G. unclassified, F. Erysipelotrichaceae* on endometrioid cancer using pleiotropy-robust methods (Supplementary Table 9 and Supplementary Figure 1) were inconsistent in magnitude to the IVW-derived and original Wald ratio estimate. Specifically, the MR-Egger effect estimate was in the same direction and both the weighted median and weighted mode effect estimates were in the opposite direction to the IVW, with all CIs spanning the null (MR-Egger OR: 1.01; 95% CI: 0.70, 1.46; P=0.97, weighted median OR: 0.998; 95% CI: 0.92, 1.08; P=0.96 and weighted mode OR: 0.99; 95% CI: 0.86, 1.13; P=0.85). Given the inconsistencies between the originally observed causal effect estimates (derived by both Wald ratio and IVW methods) and pleiotropic-robust methods, and the magnitude of the effect estimates, coupled with the phenotypic associations with the original single SNP associated with *G. unclassified, F. Erysipelotrichaceae* used in the main analysis, it is possible that the apparent causal effect reflects violations of key MR assumptions.

#### Colocalisation

To determine whether the single genetic variant used as an instrument for *G. unclassified, F. Erysipelotrichaceae* was associated with variation in both this microbial trait and endometrioid cancer, which is necessary but not sufficient for establishing causality, we performed genetic colocalisation. Genome-wide data for microbial traits was only available for one of the cohorts included in the mGWAS meta-analysis, the Flemish Gut Flora Project (FGFP), which limited analyses to 2,223 participants. The SNP used to instrument *G. unclassified, F. Erysipelotrichaceae* (rs6733298) did not reach the genome-wide p-value threshold (i.e., 5×10^-8^) in this dataset alone, so the focus of these analyses was the difference between tested hypotheses. The colocalisation results (Table 2) firstly showed that the genetic variant was more strongly related to *G. unclassified, F. Erysipelotrichaceae* than to endometrioid cancer (posterior probabilities for H1 and H2 were 0.51 and 0.06, respectively) and that it was unlikely that neither trait had a related causal variant in the region (posterior probability for H0=0.29). There was little evidence that *G. unclassified, F. Erysipelotrichaceae* and endometrioid cancer risk shared a causal variant (i.e., the posterior probability for H4=0.04). There was also overall weak evidence that the observed causal relationship was driven by linkage disequilibrium (LD) (posterior probability of H3=0.11), though this probability was larger than there being a shared causal variant. Regional association plots confirmed these findings (Figure 2), showing that, whilst the variant was associated with the microbial trait, there was little evidence for an association of this variant (or variants in the surrounding genomic region) with endometrioid cancer risk. Whilst there was an additional peak showing another SNP associated with endometrioid cancer, approximately 500kb downstream of the microbiome-related SNP, there was no evidence that this was in LD with the SNP related to the microbial trait (Figure 2).

**Figure 2.**
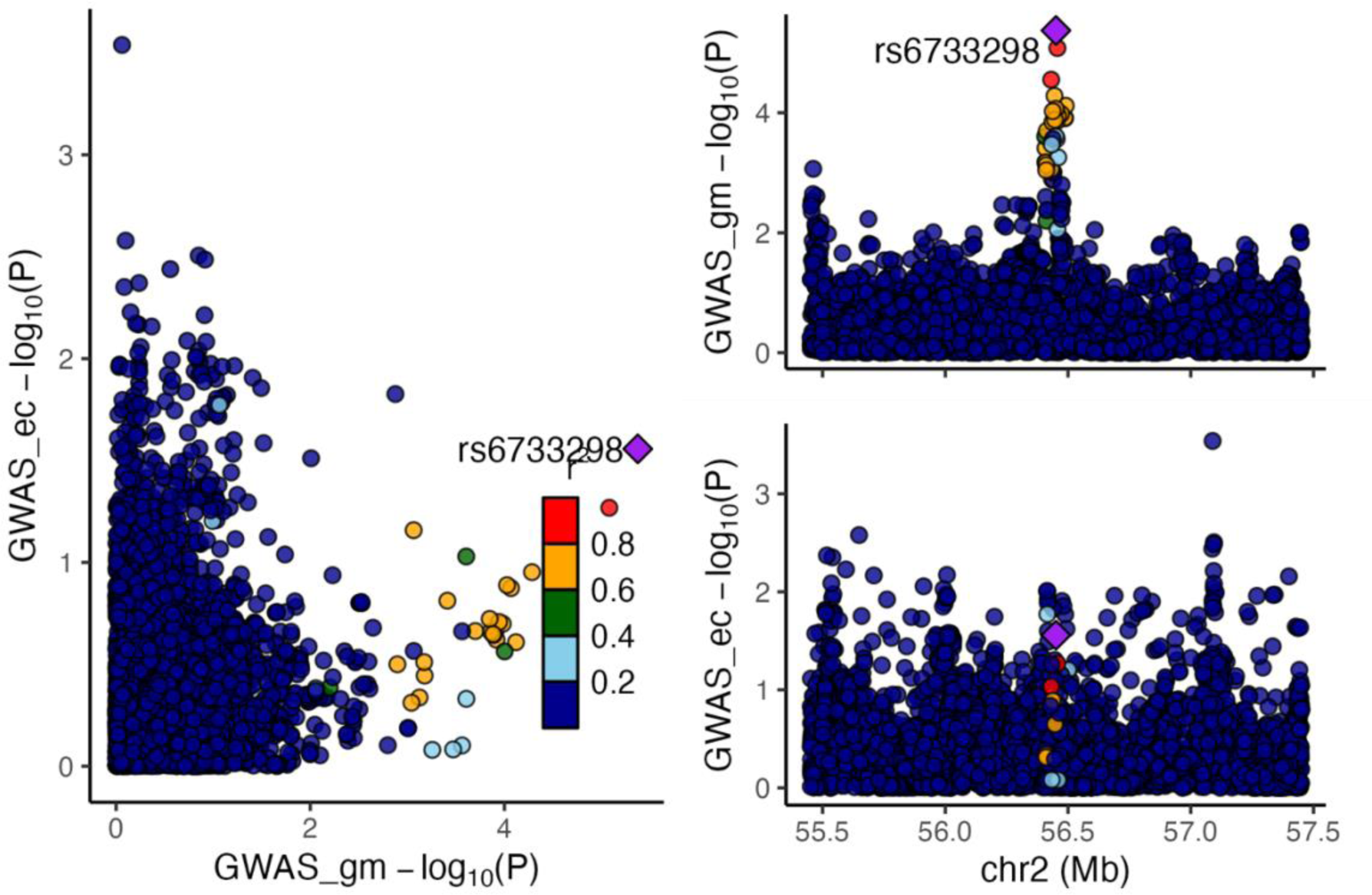
Colocalisation results for lead SNP (rs6733298) associated with the *G. unclassified, F, Erysipelotrichaceae* P/A microbial trait with endometrioid cancer. *Abbreviations: AB = abundance; ec = endometrial cancer; FGFP = Flemish Gut Flora Project; GM = gut microbiome; GWAS = genome-wide association study. Regional association plots, generated from LocusCompareR, showing the −log10(P-value) where the lead SNP (rs6733298) associated with the G. unclassified, F. Erysipelotrichaceae P/A microbial trait is represented by a purple diamond. These plots were created using the FGFP and O’Mara et al.* (*2018*) *full summary-level data for microbial traits and endometrioid cancer, respectively*.

**Table 2.** Posterior probabilities for colocalisation analyses relating to the association between G. unclassified, F, Erysipelotrichaceae P/A and endometrioid cancer.

| Posterior probabilities | H0 | H1 | H2 | H3 | H4 |
| --- | --- | --- | --- | --- | --- |
| G. unclassified, F. Erysipelotrichaceae (P/A) | 0.29 | 0.51 | 0.06 | 0.11 | 0.04 |

*Abbreviations: AB = abundance; P/A = presence vs absence. Letters in the microbial trait name represent the taxon classification level from which that microbial trait was observed, with “G” and “F” representing “genus” and “family”, respectively. All microbial traits that were not confidently classified at the genus level were organised into unclassified groups within higher classification levels (represented by “unclassified”). H0: Neither trait has a genetic association in the region, H1: Only the first trait (i.e., the microbial trait) has a genetic association in the region, H2: Only the second trait (i.e., endometrioid cancer risk) has a genetic association in the region, H3: Both traits are associated but have different causal variants and H4: Both traits are associated and have the same causal variant.*

#### Reverse MR

To finally examine whether the causal effect observed in the main analyses was driven by reverse causation (i.e., where the SNP used as an instrument for the microbial trait was affecting endometrioid cancer and this, in turn, had an effect on the microbial trait), two-sample MR analyses in the reverse direction were performed. This analysis aimed to assess whether endometrioid cancer had a causal effect on the presence of *G. unclassified, F. Erysipelotrichaceae*. Instruments for endometrioid cancer were obtained from a recent large GWAS on endometrial cancer, where a total of 10 SNPs were used as instruments for endometrioid cancer. The results showed little evidence that endometrioid cancer had a strong effect on the presence of *G. unclassified, F. Erysipelotrichaceae* (OR: 0.97; 95% CI: 0.66, 1.43; P=0.89) (Supplementary Table 10).

## DISCUSSION

We used two-sample MR to assess the causal effect of 14 microbial traits on endometrial cancer, using a comprehensive analysis pipeline previously applied in a similar context (18). We initially found that an unclassified group of bacteria in the *Erysipelotrichaceae* family was likely to increase the risk of endometrioid cancer; however, sensitivity analyses suggested that there were likely violations of core MR assumptions, specifically the existence of horizontal pleiotropy, meaning that our initial finding is unlikely to reflect causality.

Firstly, in the manual exploration of pleiotropy we found that the SNP used as an instrument for the microbial trait of interest (rs6733298) was associated with a number of other traits with biological relevance to endometrioid cancer, including leg and body fat mass and percentage. From these analyses alone, it was not possible to tell if this indicated the presence of horizontal or vertical pleiotropy. Indeed, it could be that the relationships between rs6733298, *G. unclassified, F. Erysipelotrichaceae* and these adiposity-related traits simply reflects the causal pathway by which the gut microbiome influences endometrial cancer. However, it is equally plausible that rs6733298 independently influences both this gut microbial trait and adiposity, where the latter only influences endometrial cancer. By increasing our p-value threshold to allow more instruments to be used to proxy the microbial trait, we were able to conduct pleiotropy-robust methods. The results of these methods were inconsistent, whereby the estimates derived from the weighted median and mode methods were inconsistent to estimates derived using the Wald ratio, IVW and MR-Egger methods. The weighted median estimator assumes that no more than 50% of the instruments are invalid and the weighted mode estimator assumes that the greatest number of similar estimates are derived from valid instruments. Thus, the inconsistency between these two methods and all other methods undertaken may indicate that the more lenient p-value threshold in this scenario enabled the inclusion of mostly invalid SNPs, even when restricting to those that had directionally consistent effect estimates across the studies included in the mGWAS. This further highlights the requirement to use SNPs that are robustly and consistently associated with gut microbial traits in future MR analyses assessing the role of the gut microbiome in all health outcomes. Whilst the consistency between the estimates derived from the IVW and MR-Egger methods indicate that, when accounting for directionally pleiotropic SNPs (i.e., which may have in part been biasing the weighted median- and mode-based estimates), the causal effect was in the positive direction, the lack of statistical evidence for such a causal effect coupled with the findings from the manual exploration of pleiotropy, suggested that horizontal pleiotropy may very likely be at play. Furthermore, the reliability of these methods and their ability to derive informative estimates suffer with increasingly limited numbers of independent SNPs, which is the case in this context, where only four SNPs were available for MR analyses. Additionally, the colocalisation results showed that there was no shared causal variant affecting both the microbial trait and endometrial cancers and that the observed relationship was likely not driven by LD. Given that colocalisation is a requirement for causality, these analyses coupled with other sensitivity analyses undertaken, suggest our initial finding was not reflective of a causal relationship. Lastly, given that the SNP used as an instrument for the microbial trait in the main analyses was more strongly associated with the microbial trait compared to the endometrioid cancer outcome and the reverse MR analyses found little evidence for a reverse effect of endometrioid cancer on *G. unclassified, F. Erysipelotrichaceae*, the main analyses were likely not biased by reverse causation (i.e., where the SNP primarily influences the outcome, which, in turn, influences the exposure). This work therefore demonstrates the importance of conducting sensitivity analyses to assess the robustness of MR findings.

A particular strength of this study is the methods used for selecting SNPs associated with the gut microbiome and the comprehensive sensitivity analyses applied to test the robustness of main analyses. Previous MR studies assessing the causal role of gut microbial traits on other diseases or traits have used a lenient p-value threshold and these signals have not been replicated in subsequent studies (19). Our analysis highlights that this is not an appropriate strategy, as it likely leads to the inclusion of invalid SNPs, biased results and inaccurate conclusions. The mGWAS used in this analysis improved on this as it was a meta-analysis of multiple studies and only selected instruments that reached a genome-wide significance (p<5×10^-08^). Additionally, we used one of the largest endometrial cancer GWASs to date, so were 88% powered to detect an OR of 1.3 in this MR analysis. This means, however, that we may have missed other smaller effect estimates between different microbial traits and endometrial cancer.

There are a number of limitations of this work, which relate to the assumptions of MR. Firstly, there cannot be any pathway between the genetic instruments used to proxy the exposure (here, the microbial traits) and the outcome of interest (here, endometrial cancer) that is independent of the exposure. Given that the biological mechanisms by which host genetic variation relates to the gut microbiome are poorly understood, we cannot rule out that the microbiome GWAS signals are due to unaccounted upstream factors in the host (and we are therefore observing reverse causation in MR analyses) or there may be horizontal pleiotropy at play (i.e., independent associations between genetic variants used as instrumental variables and endometrial cancer). Therefore, further work in the field, with harmonised and large-scale datasets, is required to better understand the mechanisms behind how host genetic variation could drive variation in the microbiome. Secondly, our colocalisation analyses were limited by power, as we were only able to use the full genome-wide summary statistics from the FGFP data alone. However, these results are still a helpful illustration that there appears to be no variant associated with both the gut microbiome and endometrioid cancer in this region. Relating to the availability of full genome-wide summary statistics, the reverse MR analyses may also have been underpowered.

Another issue with any potential findings of this work is the taxonomical classification of each microbial trait. For the relationship we observed in the original MR analyses, we found that an unclassified group of bacteria in the *Erysipelotrichaceae* family had a causal effect on endometrioid cancer. This is not particularly useful information if we cannot pinpoint the group of bacteria that is responsible for this effect and could be a possible target for potential interventions (if, indeed, the estimate did reflect a causal effect). Going forward, we would need more specific classification of bacterial groups in order to fully understand any causal relationships and be able to inform preventative strategies to reduce the risk of endometrial cancer.

This study initially found evidence for a causal effect of an unclassified group of bacteria in the *Erysipelotrichaceae* family and increased endometrioid cancer risk. However, subsequent sensitivity analyses suggested this finding may be due to alternative factors independent of the causal pathway. Whilst there were some limitations with this work, this study, along with previous similar work (18), has been useful in highlighting the need for stringent analysis pipelines when conducting MR studies.

## METHODS

### Study design

Two-sample MR was used to investigate the causal relationship between different traits of the gut microbiome and endometrial cancer. SNPs associated with the exposure and outcome of interest were obtained from summary data of two independent GWASs. Specifically, these data were used to estimate the causal relationship between 14 microbial traits and endometrial, endometrioid and non-endometrioid cancer risk. The Strengthening the Reporting of Observational Studies in Epidemiology using Mendelian Randomization (STROBE-MR) guidelines (20) were used to structure the reporting of this study.

### Gut microbiome GWAS data and instrument selection

SNPs associated with microbial traits were obtained from a large microbiome GWAS (mGWAS) conducted in three independent European cohorts, the Flemish Gut Flora Project (FGFP; n=2,223) and two German cohorts (FoCus (n=950) and PopGen (n=717) (10) (data available here: https://data.bris.ac.uk/data/dataset/22bqn399f9i432q56gt3wfhzlc). SNPs were identified for a total of 14 microbial traits including those that reflected a change in microbial relative abundance (AB) and the likelihood of presence vs absence (P/A) in faecal samples (Supplementary Table 1). DNA was extracted from faecal samples provided by participants and 16S rRNA gene sequencing was conducted to taxonomically classify bacteria. Following quality control (QC) steps, a total of 159 microbial traits were analysed in the mGWAS. Further information on recruitment, sampling, preparation and analysis has been previously reported (10, 21). After genotyping and QC of the FGFP cohort, 2,223 participants were used in the analyses, all from European ancestry. The FGFP cohort was used as the discovery cohort in the analysis, and all SNPs that reached an association test p-value threshold (P<1×10^−05^) in this cohort were taken forward into a meta-analysis with the FoCus and PopGen studies. Six microbial traits were not present in these studies, so the number of microbial traits in the meta-analysis was 153. The meta-analysis was performed using the inverse variance fixed effects method and SNPs were considered “meta-supported” if they reached a smaller p-value of association in the meta-analysis than in the FGFP cohort alone. The genome-wide meta-analysis p-value threshold was defined as 2.5×10^−08^ and the study-wide p-value threshold was defined as 1.57×10^−10^, using Bonferroni correction for the number of microbial traits analysed. A total of 13 SNPs reached this genome-wide meta-analysis p-value, each associated with an independent microbial trait, were selected as genetic instruments, where two SNPs reached the study-wide p-value threshold and 11 SNPs reached the genome-wide threshold. One additional SNP was also selected, as it was the most consistently previously reported SNP associated with bacteria of the *Bifidobacterium* genus in the literature (22, 23). Effect estimates from the mGWAS represent an increase in the log-transformed OR for P/A microbial traits and the standard deviation (SD) change for rank normalised AB microbial traits with each effect allele carried.

### Endometrial cancer GWAS data

Data for endometrial cancer was obtained from a GWAS meta-analysis conducted using 17 studies of participants of European ancestry, totalling 12,906 endometrial cancer cases and 108,979 country-matched controls (11). Studies used in this GWAS meta-analysis included the Epidemiology of Endometrial Cancer Consortium (E2C2), the Endometrial Cancer Association Consortium (ECAC) and UK Biobank (details of studies included available in Supplementary Table 11). These analyses also distinguished between the different histological subtypes of endometrial cancer, endometrioid (cases=8,758; controls=46,126) and non-endometrioid (cases=1,230; controls=35,447). This data was downloaded from the IEU OpenGWAS (GWAS IDs: ebi-a-GCST006464-6). The OncoArray genotyping chip, containing 533,631 variants, was used to genotype 5,061 endometrial cancer cases from 10 studies. After SNP-wise QC, 469,364 SNPs remained for imputation. The 5,061 OncoArray-genotyped endometrial cancer cases were country-matched to controls. Following QC, OncoArray genotypes from 4,710 cases and 19,438 controls remained and were included in the analyses. Following SNP-wise QC, the analysis included a total of 12,906 endometrial cancer cases and 108,979 controls, which included 8,758 endometrioid cases and 1,230 non-endometrioid cases. The remaining cases were either of mixed histology or the study did not include information about histology. Per-allele ORs were computed using logistic regression and estimated ORs from different studies were combined in a fixed effects inverse variance weighted meta-analysis. A conventional genome-wide p-value threshold (p<5×10^-08^) was used to select SNPs. Further details on the studies used, genotyping, QC and analyses has previously been published (11). This meta-analysis confirmed seven of the eight previously published loci associated with endometrial cancer and nine further independent risk loci were identified including eight newly reported regions and a locus that had previously been identified by a joint endometrial-colorectal cancer analysis. The seven previously published variants and three of the newly identified variants also reached the genome-wide p-value threshold in an analysis restricted to endometrioid cancer. No SNPs reached the genome-wide p-value threshold when analyses were restricted to non-endometrioid cases.

### Statistical analyses

#### Two-sample Mendelian randomization

In these analyses, two-sample MR was used using the TwoSampleMR package (version 0.5.6) (24) in R (version 4.1.3) to examine the causal relationship between 14 different microbial traits and endometrial cancer (both overall and subtype-specific).

The methods used here have previously been described (18). In brief, summary-level data for each of the 14 microbial trait-associated SNPs were extracted from both the mGWAS and the endometrial cancer GWAS meta-analysis. If a particular SNP wasn’t present in the endometrial cancer data, a proxy SNP was used that was correlated with the original SNP (i.e., r^2^ >0.8). The proportion of variance in each microbial trait explained by each SNP (R^2^) and the strength of the instrument (F-statistic) has previously been calculated (18) (Supplementary Table 1). For binary microbial traits (i.e., presence vs absence), the R^2^ was calculated using the “get_r_from_lor” function of the TwoSampleMR package (24) and this estimate was then squared. For continuous traits (i.e., abundances) the R^2^ was calculated using the following formulas (25):

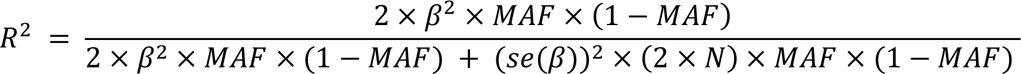

Where β is the effect size for the association between a given SNP and the exposure, MAF is the minor allele frequency, se(β) is the corresponding standard error of the effect size, and N is the sample size used to estimate the SNP-exposure association.

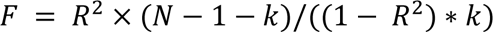

Where R^2^ is the proportion of variance in the exposure explained by the instrument, N is the sample size used to estimate the SNP-exposure association, k is the number of SNPs included in the instrument.

The exposure and outcome datasets were harmonised so that the effect of each SNP on the exposure and the effect of that same SNP on the outcome corresponded to the same allele. Palindromic SNPs (i.e., where the effect/alternative allele are either A/T or G/C combinations) were harmonised, where possible, using the allele frequencies and excluded if it was not possible to harmonise them (i.e., when the MAF of the SNP was above 0.42).

As there was only one SNP associated with each microbial trait, the Wald ratio method was used to estimate the effect of each microbial trait on endometrial cancer. This was calculated by dividing the SNP-outcome association by the SNP-exposure association (8).

The effect estimates obtained from two-sample MR analyses represent the OR for endometrial cancer risk with either each SD higher relative abundance (AB) of each continuous microbial trait or an approximate doubling of the genetic liability to the presence vs absence (P/A) of each binary microbial trait. P-values were used as a continuous indication of the strength of an association, where the P-value and beta-coefficient were the basis of conclusions drawn from analyses. Given the relative nature of microbial traits, there was no correction for multiple testing.

### Sensitivity analyses

#### Assumptions of MR analyses

There are three core assumptions that must be met when using MR to estimate a causal relationship between exposure and outcome (Figure 3). The first is that the genetic instruments used to proxy the exposure are robustly associated with this exposure. The second is that there are no confounders of the genetic instruments used to proxy the exposure and the outcome (i.e., confounders driven by population substructure, assortative mating or intergenerational effects). Finally, the third assumption is that the genetic instruments used to proxy the exposure are only associated with the outcome through the exposure (9, 26, 27). As SNPs used were associated with the exposure in all cases and both data sources were of European descent, a series of sensitivity analyses were conducted to test for violations of, predominantly, the third MR assumption.

**Figure 3.**
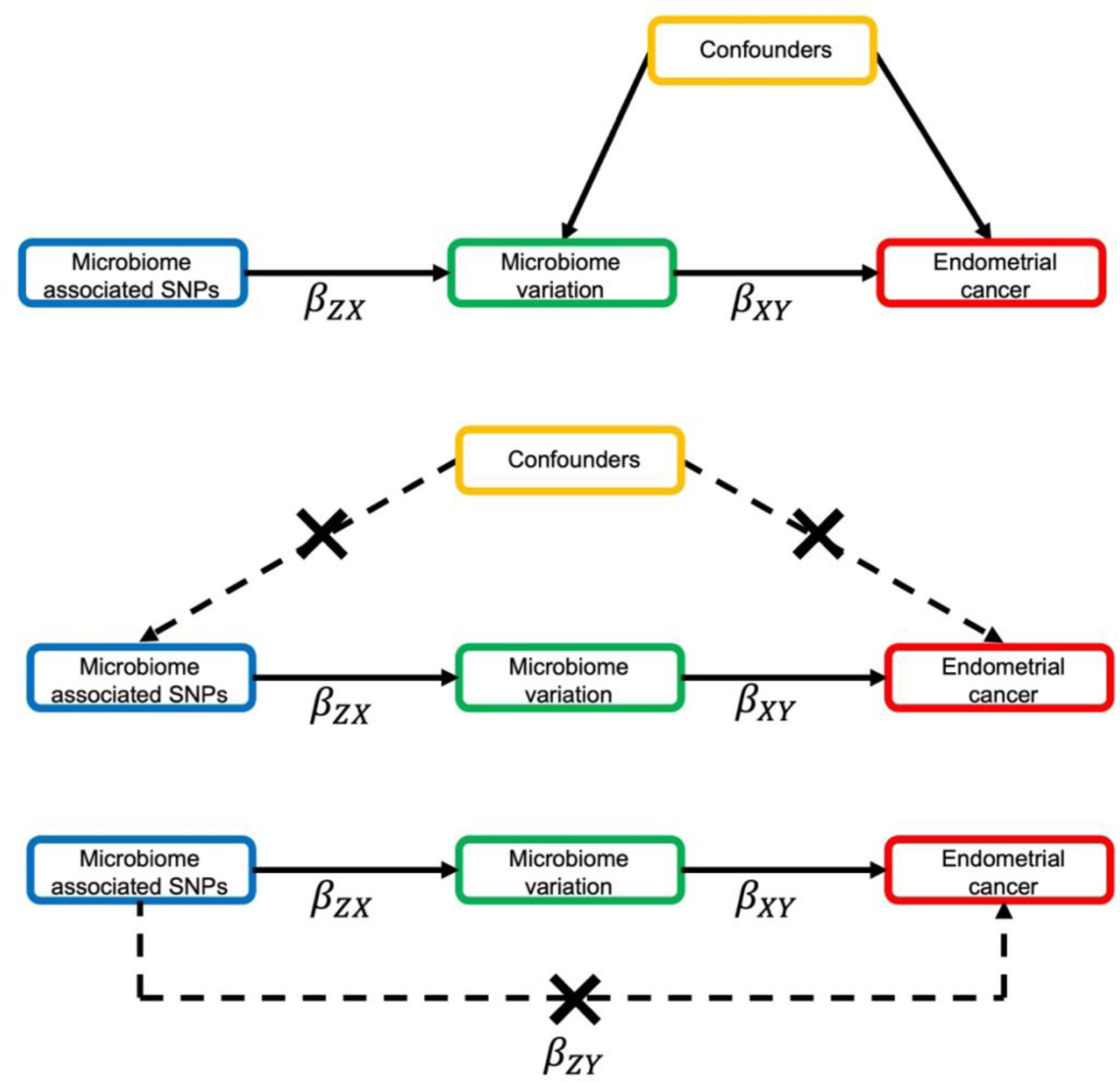
MR framework applied to assess the causal effect of the gut microbiome on endometrial cancer risk. Abbreviations: MR = Mendelian randomization; SNP = single nucleotide polymorphism. MR aims to improve causal inference by reducing the limitations of traditional observational epidemiology studies and relies on three key assumptions: (A) the SNPs are strongly associated with the exposure (the gut microbiome); (B) there is no confounding between the SNPs, used to proxy the exposure, and the outcome (endometrial cancer); and (C) the SNPs are not independently associated with the outcome other than through the exposure. When these assumptions are met, microbiome-associated genetic variants can be used as instruments to assess the causal effect of gut microbial traits on endometrial cancer. In two-sample MR, the SNP-outcome (β_ZY_) and the SNP-exposure (β_ZX_) effect estimates are derived from two independent samples and the causal effect of the exposure on the outcome (β_XY_) calculated as a ratio of β_ZY_ and β_ZX_.

#### Manual exploration of pleiotropy

Since there were only singular instruments for each microbial trait, pleiotropy-robust methods that usually require multiple genetic variants were not possible to apply within this context. Therefore, to assess whether any causal relationships found between microbial traits and endometrial cancer were affected by horizontal pleiotropy, the SNPs being used as instruments were looked up in the IEU OpenGWAS (15, 24) and PhenoScanner (16) platforms to determine if these SNPs had previously been reported to be associated with either endometrial cancer or traits associated with endometrial cancer risk, which could be independent from the gut microbiome. Based on the number of results returned by PhenoScanner and the number of phenotypes in the IEU OpenGWAS platform, a lenient p-value threshold (P<1×10^-4^) was set as the multiple testing threshold to define the presence of associations between the SNPs and traits.

#### Two-sample MR using a lenient p-value threshold

When using the traditional p-value threshold for instrument selection (p<5×10^-8^), there was only one SNP associated with each microbial trait; therefore, it was not possible to conduct formal pleiotropy-robust methods that require multiple genetic instruments. To increase the number of SNPs associated with each microbial trait (and thus enable the application of these pleiotropy-robust methods), a more lenient p-value threshold (p<1×10^-5^) was used for instrument selection and SNPs that met this threshold were then restricted to those with directionally consistent effect estimates between the three studies in the mGWAS (10). These SNPs were obtained from previously published work (18). The inverse-variance weighted (IVW) method was used as the main analysis, which meta-analyses effect estimates obtained by the Wald ratio across SNPs, weighted by the inverse variance of the SNP-outcome association using fixed effects. As using a more lenient p-value threshold increases the risk of weak instrument bias and horizontal pleiotropy, further methods were applied to test for this. The weighted median (28), weighted mode (29) and MR-Egger (30) regression methods were used and effect estimates were compared across all methods.

The weighted median (28) method can produce a consistent causal estimate even when up to 50% of SNPs are invalid instruments. Weighting the individual SNP effect estimates by the inverse variance of the SNP-outcome association, the method then orders the weighted effect estimates by magnitude and selects the median value as the causal effect estimate. Similarly, assuming that the most common causal effect estimate is the true causal effect (even if the majority of instruments are invalid), the weighted mode method clusters the weighted effect estimates of each SNP by similarity and then estimates the causal effect based on the largest group of SNPs (29). The MR-Egger (30) method estimates the causal effect adjusted for any directional pleiotropy, by allowing a non-zero intercept in the relationship between SNP-outcome and SNP-exposure associations, where the intercept provides an indication of the presence and magnitude of directional pleiotropy.

#### Colocalisation

Colocalisation analyses were performed using the ‘coloc’ package in R (as previously described (18)) using the default parameters (i.e., with the prior probabilities of the SNP being associated with the exposure, the outcome or both traits being specified as 1×10^-04^, 1×10^-04^ and 1×10^-05^, respectively (31)), additionally specifying that the exposures (i.e., microbial traits) were binary or continuous, where appropriate (i.e., depending on whether the trait was AB or P/A) and that the outcome (i.e., endometrioid cancer risk) was binary with the ‘quant’ or ‘cc’ option, respectively). Bayes factor computation was used to generate 5 posterior probabilities (H0-H4) characterised by the following outcomes: (H0) neither trait has a genetic association in the region; (H1) only the microbial trait of interest has a genetic association in the region; (H2) only endometrial cancer has a genetic association in the region; (H3) both traits are associated but have different causal variants and (H4) both traits are associated and have the same causal variant. We used a posterior probability threshold ≥0.80 to indicate evidence of a shared common causal variant between microbial traits and endometrioid cancer. Full summary statistics were only available for the FGFP cohort for gut microbiome variation; therefore, genetic variants ±1Mb of the lead SNP associated with microbial traits for which there was evidence for a causal impact on endometrial cancer in our main analyses were extracted from the FGFP and E2C2 genome-wide datasets. Regional association plots were generated to visualise genetic colocalisation using the LocusCompareR package (32).

#### Reverse two-sample MR

To determine if any association found in the main analyses was explained by reverse causation (i.e., where the SNP associated with the microbial trait was actually predominantly related to endometrial cancer, which was then causing a change in the microbial trait), two-sample MR in the reverse direction was conducted (i.e., where endometrial cancer was treated as the exposure and the microbial traits for which there was evidence for an effect on endometrial cancer in the main analysis was considered the outcome). SNPs associated with endometrial cancer were obtained from a large GWAS meta-analysis of 12,906 cases and 108,979 controls (11). SNPs were selected by using a conventional genome-wide p-value threshold (p<5×10^-8^). For endometrial cancer (i.e., all histologies), 16 SNPs were available as instruments. For endometrioid cancer, 10 SNPs were available instruments. No SNPs reached the genome-wide p-value threshold for non-endometrioid cancer; therefore, reverse two-sample MR analyses were not possible to conduct. We extracted SNPs associated with the endometrial cancer outcomes from the FGFP cohort (10) (as full summary statistics were only available for the this cohort) for any microbial traits that had been found to have a causal effect on endometrial cancer in the main analyses. MR analyses were conducted as above and, as multiple SNPs were available for each exposure, the IVW method was used with comparison to pleiotropy-robust methods (i.e., weighted median, weighted mode and MR-egger methods) described above.

## Supporting information

STROBE-MR checklist

## Author contributions

KHW and CH conceived the study; KHW and CH provided supervision and training; EF performed the data analysis and prepared original manuscript. All authors critically reviewed the manuscript and contributed important intellectual content.

## Data availability statement

GWAS summary-level data used in this study were publicly available for microbiome GWAS (mGWAS) conducted by Hughes et al (10) (available here: https://data.bris.ac.uk/data/dataset/22bqn399f9i432q56gt3wfhzlc). We used the full meta-analysis results (i.e., listed as sub-level “method_em”) for the main analysis and FGFP-only analysis (i.e., listed as sub-level “method_score”) for the sensitivity analyses, including reverse MR and colocalisation analyses. Summary-level data for overall and subtype specific endometrial cancer was publically available on the IEU OpenGWAS (GWAS IDs: ebi-a-GCST006464-6).

## Data Availability

GWAS summary-level data used in this study were publicly available for microbiome GWAS (mGWAS) conducted by Hughes et al (10) (available here: https://data.bris.ac.uk/data/dataset/22bqn399f9i432q56gt3wfhzlc). We used the full meta-analysis results (i.e., listed as sub-level method_em) for the main analysis and FGFP-only analysis (i.e., listed as sub-level method_score) for the sensitivity analyses, including reverse MR and colocalisation analyses. Summary-level data for overall and subtype specific endometrial cancer was publically available on the IEU OpenGWAS (GWAS IDs: ebi-a-GCST006464-6).

https://gwas.mrcieu.ac.uk/datasets/ebi-a-GCST006464/

https://gwas.mrcieu.ac.uk/datasets/ebi-a-GCST006465/

https://gwas.mrcieu.ac.uk/datasets/ebi-a-GCST006466/

