## Supplementary Figures for "Exploring the causal role of the human gut microbiome in endometrial cancer: a Mendelian randomization approach"

**Figure 1: Forward MR results for effect of the presence or absence of *G. unclassified, F. Erysipelotrichaceae* on endometrioid cancer risk using a more lenient p-value threshold.**

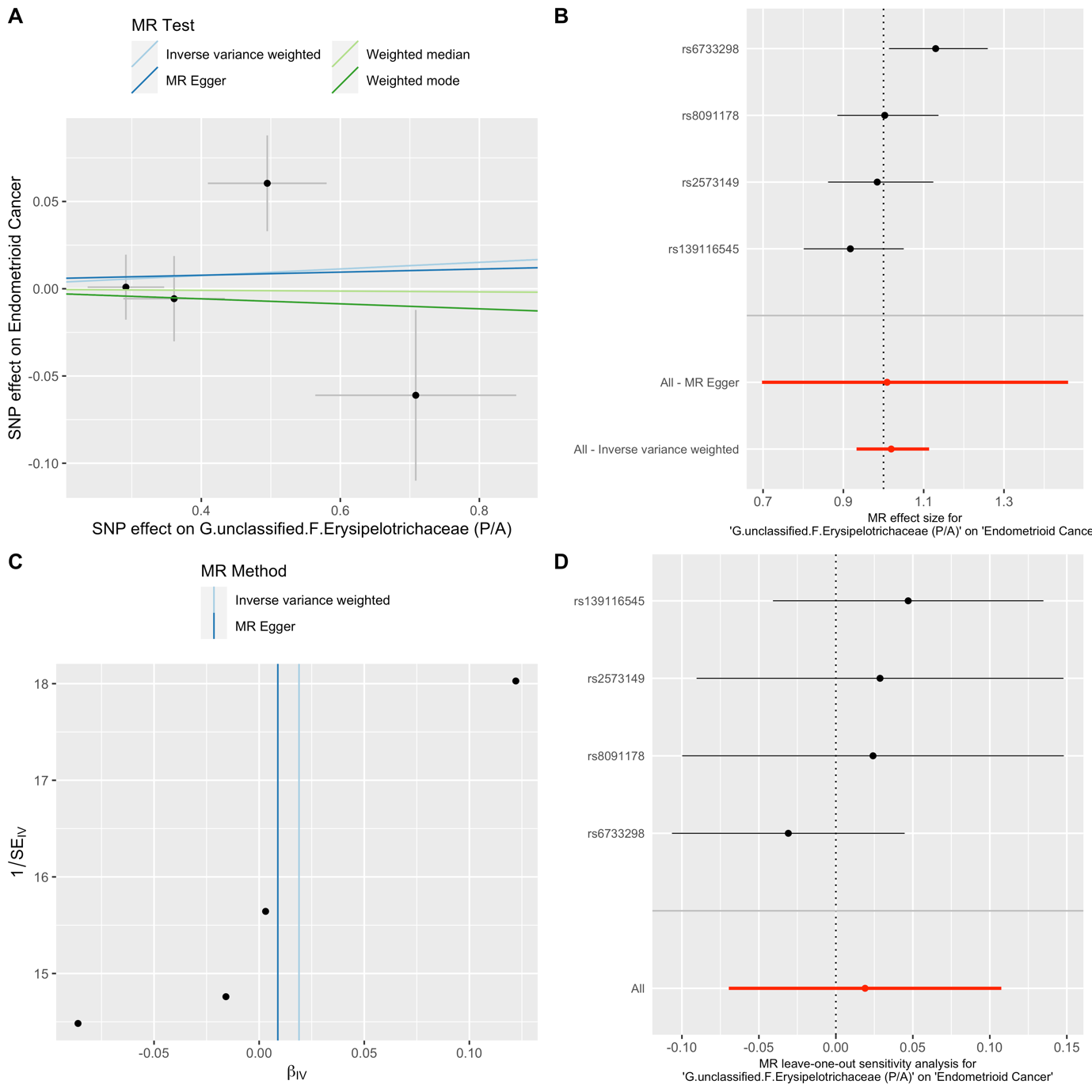

*Abbreviations: MR = Mendelian randomization; SNP = single nucleotide polymorphism; P/A = presence versus absence. These plots were generated using the Two Sample MR package, showing the results of reverse MR analyses using a lenient p-value threshold (P<1x10-4) (enabling 4 SNPs to be used as instrumental variables for G. unclassified, F. Erysipelotrichaceae (P/A) and therefore pleiotropy-robust methods to be used) to test the causal effect of the presence versus absence of this microbial trait on endometrioid cancer. A) scatter plot to compare all four methods; B) forest plot comparing effect of individual SNPs with the inverse variance weighted and MR Egger estimates; C) funnel plot to check for asymmetry; D) leave one out analysis to check if any one SNP is driving pleiotropy or asymmetry in the estimate.*
