## Supplementary material for "Exploring the causal role of the human gut microbiome in endometrial cancer: a Mendelian randomization approach": STROBE-MR checklist

**STROBE-MR checklist of recommended items to address in reports of Mendelian randomization studies**^1^ ^2^

| **Item No.** | **Section** | **Checklist item** | **Page No.** | **Relevant text from manuscript** |
| --- | --- | --- | --- | --- |
| 1 | **TITLE and ABSTRACT** | Indicate Mendelian randomization (MR) as the study’s design in the title and/or the abstract if that is a main purpose of the study | 1-2 | TITLE: Exploring the causal role of the human gut microbiome in endometrial cancer: a Mendelian randomization approach, the abstract also details this |
|  | **INTRODUCTION** |  |  |  |
| 2 | **Background** | Explain the scientific background and rationale for the reported study. What is the exposure? Is a potential causal relationship between exposure and outcome plausible? Justify why MR is a helpful method to address the study question | 3-4 | The background and rationale of the study is detailed in the first three pages of introduction, justification for using MR is explained in first paragraph on page 4 |
| 3 | **Objectives** | State specific objectives clearly, including pre-specified causal hypotheses (if any). State that MR is a method that, under specific assumptions, intends to estimate causal effects | 4 | The second paragraph on page 4 clearly states objectives of the study |
|  | **METHODS** |  | 10-17 |  |
| 4 | **Study design and data sources** | Present key elements of the study design early in the article. Consider including a table listing sources of data for all phases of the study. For each data source contributing to the analysis, describe the following: | 11-13 | The first three paragraphs of the methods detail study design and the data sources used |
|  | a) | Setting: Describe the study design and the underlying population, if possible. Describe the setting, locations, and relevant dates, including periods of recruitment, exposure, follow-up, and data collection, when available. | 11-13 | Details of study design and populations included in the study are detailed in the first three paragraphs of the methods, there is also additional information on studies included in the outcome dataset in Supplementary Table 11 |
|  | b) | Participants: Give the eligibility criteria, and the sources and methods of selection of participants. Report the sample size, and whether any power or sample size calculations were carried out prior to the main analysis | 10-13 | Details of participants and sample size on page 12-13. Power calculations on page 10 |
|  | c) | Describe measurement, quality control and selection of genetic variants | 12-13 | Details of measurement and QC and selection of genetic variants on pages 12-13 |
|  | d) | For each exposure, outcome, and other relevant variables, describe methods of assessment and diagnostic criteria for diseases | 12-13 | More detailed information on this is available in the original publications which are referenced within the text |
|  | e) | Provide details of ethics committee approval and participant informed consent, if relevant | 12-13 | This information is available in the original publications of the datasets used, which are referenced within the text |
| 5 | **Assumptions** | Explicitly state the three core IV assumptions for the main analysis (relevance, independence and exclusion restriction) as well assumptions for any additional or sensitivity analysis | 15, 9 | The last paragraph on page 15 clearly details the assumptions of MR. Details of assumptions of sensitivity analyses are provided in the discussion on page 9 |
| 6 | **Statistical methods: main analysis** | Describe statistical methods and statistics used | 14-16 |  |
|  | a) | Describe how quantitative variables were handled in the analyses (i.e., scale, units, model) | 15 | Paragraph 3 of page 15 |
|  | b) | Describe how genetic variants were handled in the analyses and, if applicable, how their weights were selected | 14-16 | Weights were not provided but the methodology for MR effect estimation is provided |
|  | c) | Describe the MR estimator (e.g. two-stage least squares, Wald ratio) and related statistics. Detail the included covariates and, in case of two-sample MR, whether the same covariate set was used for adjustment in the two samples | 14-16 | Methodology for Wald ration and IVW are provided as well as how R-squared and F statistic were calculated |
|  | d) | Explain how missing data were addressed |  | n/a |
|  | e) | If applicable, indicate how multiple testing was addressed | 15 | “Given the relative nature of microbial traits, there was no correction for multiple testing” |
| 7 | **Assessment of assumptions** | Describe any methods or prior knowledge used to assess the assumptions or justify their validity | 14 | F-statistic and R-squared calculations are provided (relating to the “relevance” assumption). |
| 8 | **Sensitivity analyses and additional analyses** | Describe any sensitivity analyses or additional analyses performed (e.g. comparison of effect estimates from different approaches, independent replication, bias analytic techniques, validation of instruments, simulations) | 15-17 | Sensitivity analyses used described on pages 15-17 |
| 9 | **Software and pre-registration** |  |  |  |
|  | a) | Name statistical software and package(s), including version and settings used | 12-18 | All packages used described |
|  | b) | State whether the study protocol and details were pre-registered (as well as when and where) | n/a | n/a |
|  | **RESULTS** |  |  |  |
| 10 | **Descriptive data** |  | 4-8 |  |
|  | a) | Report the numbers of individuals at each stage of included studies and reasons for exclusion. Consider use of a flow diagram | 4, 12-13 | Details of sample size in first paragraph of methods as well as in the methods page 12-13 |
|  | b) | Report summary statistics for phenotypic exposure(s), outcome(s), and other relevant variables (e.g. means, SDs, proportions) | n/a | Not provided by individual genome-wide association studies |
|  | c) | If the data sources include meta-analyses of previous studies, provide the assessments of heterogeneity across these studies | n/a | n/a |
|  | d) | For two-sample MR:  i.  Provide justification of the similarity of the genetic variant-exposure associations between the exposure and outcome samples  ii.  Provide information on the number of individuals who overlap between the exposure and outcome studies | 4, 12-13 | Information about study population provided in methods (page 12-13) and results (page 4) |
| 11 | **Main results** |  | 4-8 |  |
|  | a) | Report the associations between genetic variant and exposure, and between genetic variant and outcome, preferably on an interpretable scale | 4 | Also in supplementary table 1 |
|  | b) | Report MR estimates of the relationship between exposure and outcome, and the measures of uncertainty from the MR analysis, on an interpretable scale, such as odds ratio or relative risk per SD difference | 5 | Also in table 1 and figure 1 |
|  | c) | If relevant, consider translating estimates of relative risk into absolute risk for a meaningful time period | n/a | n/a |
|  | d) | Consider plots to visualize results (e.g. forest plot, scatterplot of associations between genetic variants and outcome versus between genetic variants and exposure) | 24 | Figure 1 |
| 12 | **Assessment of assumptions** |  |  |  |
|  | a) | Report the assessment of the validity of the assumptions | 4-8 | F statistics provided in supplementary table 1 (assessing “relevance assumption). Various sensitivity analyses conducted to assess “exclusion restriction” assumption |
|  | b) | Report any additional statistics (e.g., assessments of heterogeneity across genetic variants, such as *I^2^*, Q statistic or E-value) | n/a |  |
| 13 | **Sensitivity analyses and additional analyses** |  |  |  |
|  | a) | Report any sensitivity analyses to assess the robustness of the main results to violations of the assumptions | 5-8 | A number of sensitivity analyses conducted |
|  | b) | Report results from other sensitivity analyses or additional analyses | 5-8 | Figure 2, supplementary table 2-10 |
|  | c) | Report any assessment of direction of causal relationship (e.g., bidirectional MR) | 8 | Supplementary table 10 |
|  | d) | When relevant, report and compare with estimates from non-MR analyses | n/a |  |
|  | e) | Consider additional plots to visualize results (e.g., leave-one-out analyses) | 6-7 | Supplementary figure 1 |
|  | **DISCUSSION** |  |  |  |
| 14 | **Key results** | Summarize key results with reference to study objectives | 8 | First paragraph of discussion |
| 15 | **Limitations** | Discuss limitations of the study, taking into account the validity of the IV assumptions, other sources of potential bias, and imprecision. Discuss both direction and magnitude of any potential bias and any efforts to address them | 9-11 | Paragraph 2, 4 and 5 of discussion |
| 16 | **Interpretation** |  |  |  |
|  | a) | Meaning: Give a cautious overall interpretation of results in the context of their limitations and in comparison with other studies | 8-11 | Comparison in paragraph 2 of page 10 |
|  | b) | Mechanism: Discuss underlying biological mechanisms that could drive a potential causal relationship between the investigated exposure and the outcome, and whether the gene-environment equivalence assumption is reasonable. Use causal language carefully, clarifying that IV estimates may provide causal effects only under certain assumptions | 11 | Limitations of biological interpretation discussed |
|  | c) | Clinical relevance: Discuss whether the results have clinical or public policy relevance, and to what extent they inform effect sizes of possible interventions | 11 | Limitations of relevance discussed |
| 17 | **Generalizability** | Discuss the generalizability of the study results (a) to other populations, (b) across other exposure periods/timings, and (c) across other levels of exposure | 10-11 | Sample size discussed |
|  | **OTHER INFORMATION** |  |  |  |
| 18 | **Funding** | Describe sources of funding and the role of funders in the present study and, if applicable, sources of funding for the databases and original study or studies on which the present study is based | 21 | Funding detailed for all authors |
| 19 | **Data and data sharing** | Provide the data used to perform all analyses or report where and how the data can be accessed, and reference these sources in the article. Provide the statistical code needed to reproduce the results in the article, or report whether the code is publicly accessible and if so, where | 21 | Data access detailed with links to publicly available datasets |
| 20 | **Conflicts of Interest** | All authors should declare all potential conflicts of interest | 21 | No conflicts of interest |

This checklist is copyrighted by the Equator Network under the Creative Commons Attribution 3.0 Unported (CC BY 3.0) license.

1.	Skrivankova VW, Richmond RC, Woolf BAR, Yarmolinsky J, Davies NM, Swanson SA, et al. Strengthening the Reporting of Observational Studies in Epidemiology using Mendelian Randomization (STROBE-MR) Statement. JAMA. 2021;under review.

2.	Skrivankova VW, Richmond RC, Woolf BAR, Davies NM, Swanson SA, VanderWeele TJ, et al. Strengthening the Reporting of Observational Studies in Epidemiology using Mendelian Randomisation (STROBE-MR): Explanation and Elaboration. BMJ. 2021;375:n2233.
